# Risk factors for mortality in pregnant women with SARS-CoV-2 infection

**DOI:** 10.1101/2020.05.31.20107276

**Authors:** Raigam J. Martinez-Portilla, Alexandros Sotiriadis, Johnatan Torres-Torres, Chatzakis Christos, Ameth Hawkins-Villarreal, Jose R. Villafan-Bernal, Rodolfo A. Gurrola-Ochoa, Francesc Figueras

## Abstract

Since the first case of pneumonia was described, SARS-CoV-2 infection (coronavirus disease [COVID]-19) rapidly spread worldwide With 94,288 infections and more than 10,000 deaths, Mexico is the third Latin-American country in number of confirmed cases and second in mortality1. A major risk factor for adverse outcome in COVID-19 infection is the presence of advance age, co-morbidities including diabetes, hypertension and obesity among other non-communicable diseases2. Epidemiological data from high-prevalence countries reveal that compared to men, women are less likely to die or to require hospital admission to intensive care. This may suggest that pregnant women are not more susceptible to infection or to experience serious complications. However, whether the presence of co-morbidities or advanced maternal age confers a higher risk of adverse outcome in pregnant women with COVID-19 is unknown3.

In this research letter, we aimed at evaluating the risk factor associated with maternal mortality secondary to COVID-19 infection in a middle-income country.

Advanced maternal age is linked to an increased risk of mortality, while diabetes is the most important risk factor for maternal death. This is partly explained by an increasing incidence of non-communicable diseases in women of advanced age which is a common feature in most countries4. In the last decades, low- and middle-income countries have experienced accelerated socio-cultural changes associated with its incorporation into the international economic community, which have increased the number of obese and diabetic population, including pregnant women5. This has caused an increased risk for complications and fatality among COVID-19 positive population2,3. Thus, policies for reducing obesity and diabetes in low- and middle-income countries are most needed to reduce the mortality of COVID-19 in pregnant women.

## Background

Since the first case of pneumonia was described, SARS-CoV-2 infection (coronavirus disease [COVID]-19) rapidly spread worldwide With 94,288 infections and more than 10,000 deaths, Mexico is the third Latin-American country in number of confirmed cases and second in mortality^1^. A major risk factor for adverse outcome in COVID-19 infection is the presence of advance age, comorbidities including diabetes, hypertension and obesity among other non-communicable diseases^2^. Epidemiological data from high-prevalence countries reveal that compared to men, women are less likely to die or to require hospital admission to intensive care. This may suggest that pregnant women are not more susceptible to infection or to experience serious complications. However, whether the presence of co-morbidities or advanced maternal age confers a higher risk of adverse outcome in pregnant women with COVID-19 is unknown^3^.

## Methods

In this prospective cohort study, data were extracted from the epidemiological surveillance system of viral respiratory diseases of Mexico(6), which includes 475 monitoring units of viral respiratory diseases, 135,116 tested population including 1,241 tested pregnant women. Inclusion criteria were pregnant women with confirmed (nasopharyngeal RT-PCR) SARS-CoV2 infection registered between January 1 and May 11, 2020. Figure 1 shows the flow diagram of the studied population. The main outcome was maternal mortality. We also recorded data on morbidities such as pregestational diabetes, chronic hypertension, pulmonary obstructive chronic disease, asthma, immunosuppression, obesity, smoking habits, chronic kidney failure, and other non-specified morbidities. Study size was not calculated, rather we included the whole national cohort. Quantitative variables were described as median and interquartile range and compared using the U-Mann-Whitney test, while categorical variables were expressed as number and percentage and compared using the Chi-squared test or Fisher’s exact test. Univariate logistic regression was used to assess the association between each predictor and maternal mortality as the main outcome. A p-value <0.05 was considered significant. STATA v.15.3 was used for statistical analyses.

**Figure 1.**
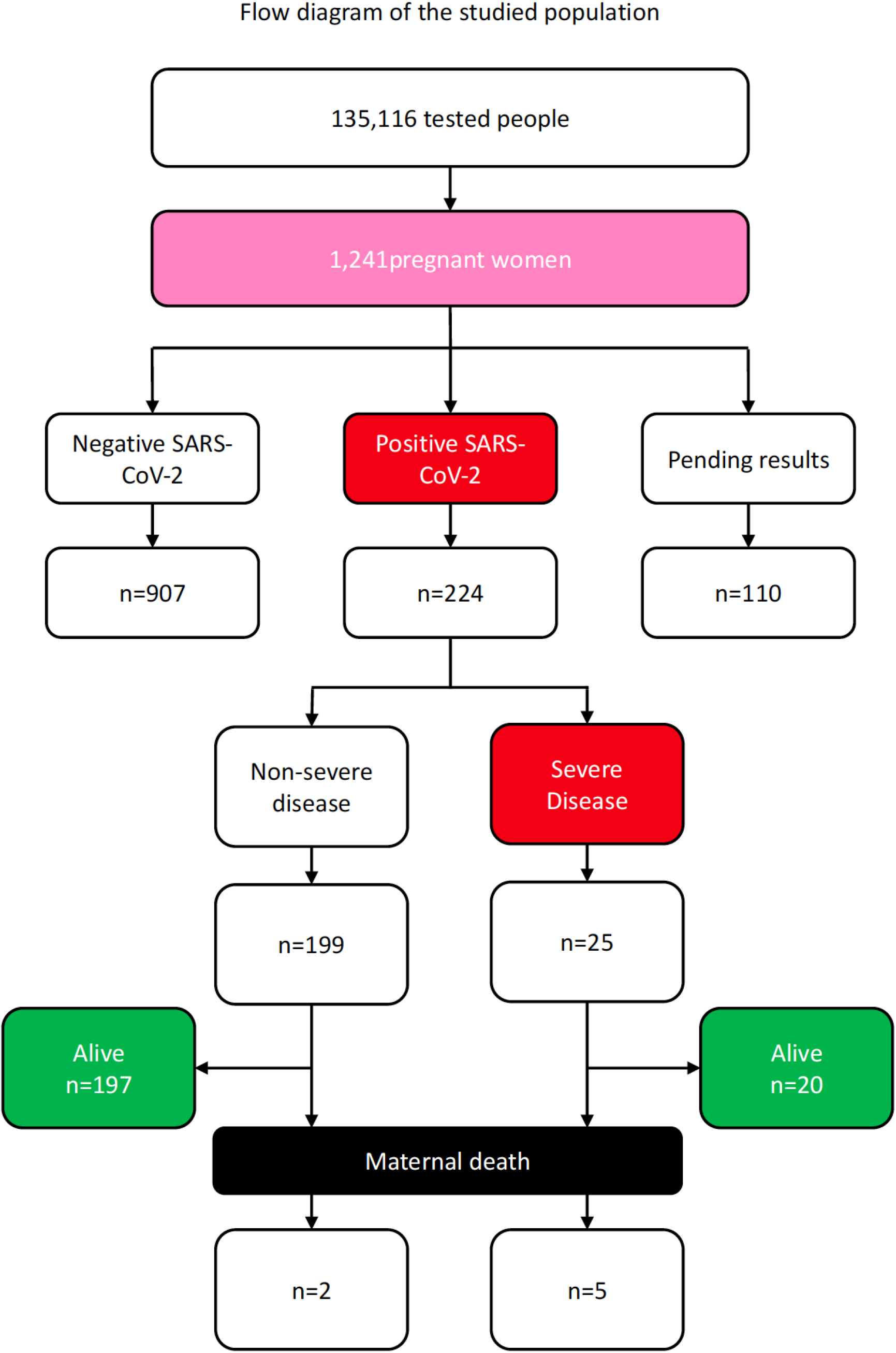
Flow diagram of the studied population

## Results

Among the 1,241 tested pregnant women, 224 (18.1%) COVID-19 were confirmed. There were 7 (3.1%) cases of maternal death. Maternal death occurred at a median of 9 days (range 4-11 days) after the onset of symptoms. Table 1 depicts the characteristics of the population by survival status. Of note, 5 out of 7 deaths occurred in women with co-morbidities. Pregnant women who died were older (median of 29 vs. 37 years; Mann-Whitney-U test p<0.001) and more likely to have co-morbidities. Figure 2 shows the odds ratios for death conferred by these epidemiological factors.

**Table 1.**
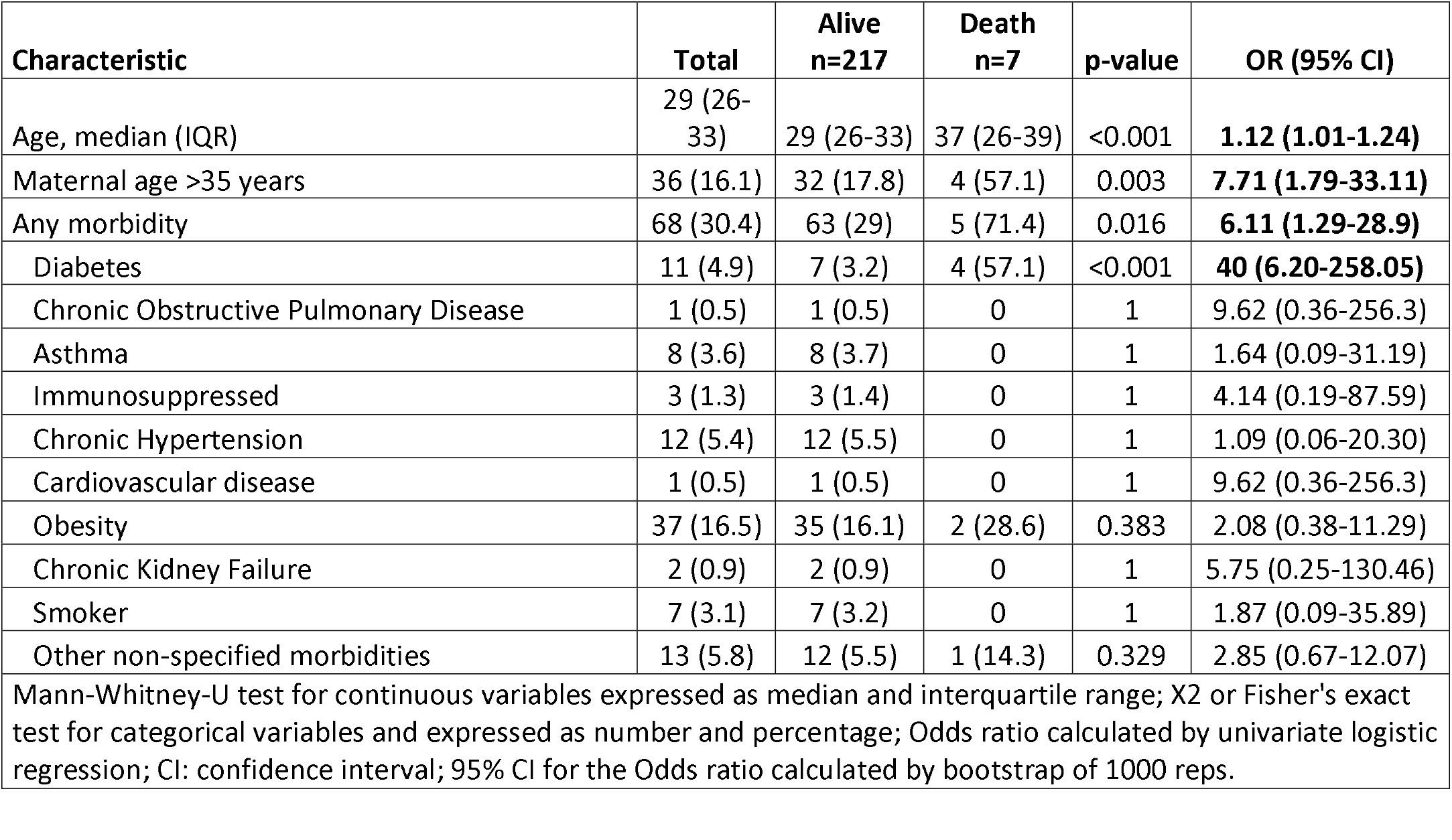
Characteristics of mortality among SARS-CoV-2 infected women.

**Figure 2.**
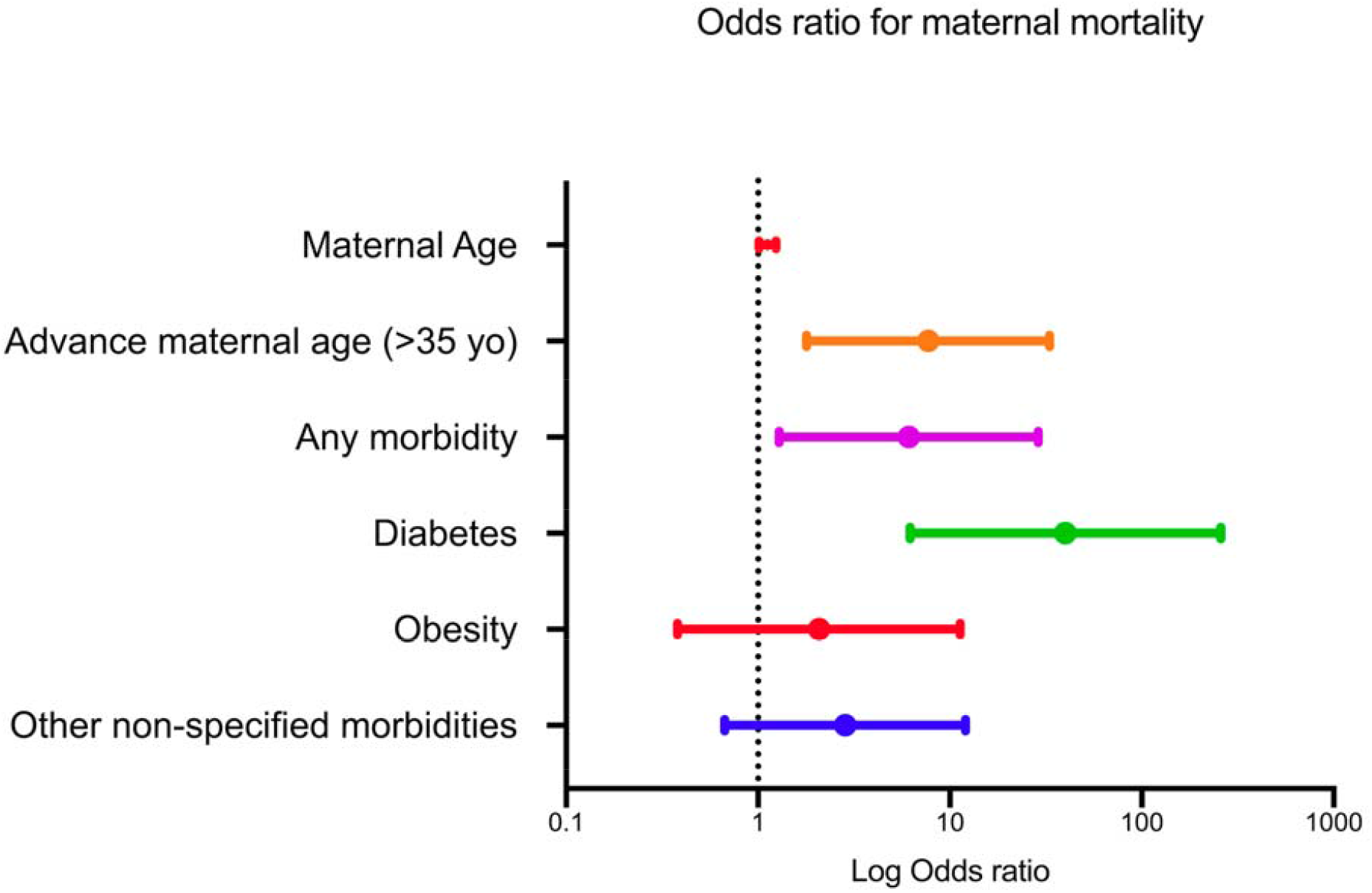
Odds ratio for maternal mortality

## Discussion

Advanced maternal age is linked to an increased risk of mortality, while diabetes is the most important risk factor for maternal death. This is partly explained by an increasing incidence of non-communicable diseases in women of advanced age which is a common feature in most countries^4^. In the last decades, low- and middle-income countries have experienced accelerated socio-cultural changes associated with its incorporation into the international economic community, which have increased the number of obese and diabetic population, including pregnant women^5^. This has caused an increased risk for complications and fatality among COVID-19 positive population^2,3^. Thus, policies for reducing obesity and diabetes in low- and middle-income countries are most needed to reduce the mortality of COVID-19 in pregnant women.

The limitation of this study is the few numbers of fatalities which does not allow more robust analyses.

In conclusion, our findings unveil that the most important risk factors for maternal mortality are advanced maternal age and diabetes. This information may lead to changes in health policies to protect most vulnerable pregnant women.

## Data Availability

Data are available by correspondence by contacting the corresponding author

